# EFCAB4B (CRACR2A) genetic variants associated with COVID-19 fatality

**DOI:** 10.1101/2022.01.17.22269412

**Authors:** Dapeng Wang, Sabina D Wiktor, Chew W Cheng, Katie J Simmons, Ashley Money, Lucia Pedicini, Asya Carlton, Alexander L Breeze, Lynn McKeown

## Abstract

The coronavirus disease 2019 (COVID-19) pandemic, caused by severe acute respiratory syndrome coronavirus 2 (SARS-CoV-2), has resulted in more than 235 million cases worldwide and 4.8 million deaths (October 2021). Severe COVID-19 is characterised in part by vascular thrombosis and a cytokine storm due to increased plasma concentrations of factors secreted from endothelial and T-cells. Here, using patient data recorded in the UK Biobank, we demonstrate the importance of variations in Rab46 (CRACR2A) with clinical outcomes. Using logistic regression analysis, we determined that three single nucleotide polymorphisms (SNPs) in the gene EFCAB4B cause missense mutations in Rab46, which are associated with COVID-19 fatality independently of risk factors. All three SNPs cause changes in amino acid residues that are highly conserved across species, indicating their importance in protein structure and function. Two SNPs, rs17836273 (A98T) and rs36030417 (H212Q), cause amino acid substitutions in important functional domains: the EF-hand and coiled-coil domain respectively. By using molecular modelling, we suggest that the substitution of threonine at position 98 causes structural changes in the EF-hand calcium binding domain. Since Rab46 is a Rab GTPase that regulates both endothelial cell secretion and T-cell signalling, these missense variations may play a role in the molecular mechanisms underlying the thrombotic and inflammatory characteristics observed in patients with severe COVID-19 outcomes.

## Introduction

Coronavirus disease 2019 (COVID-19) caused by the Severe Acute Respiratory Syndrome Coronavirus 2 (SARS-CoV-2) triggered a global pandemic in 2020 [1]. Although a disease that affects the respiratory system, increased morbidity and mortality is associated with cardiovascular complications including abnormal clotting, thrombosis and microvascular injury [3, 4]. Several studies have suggested that SARS-CoV-2 can directly infect the endothelial cells (ECs) that line the blood vessels: indeed endothelial dysfunction is found in severe cases of COVID-19 [5-7]. Activation of ECs triggers exocytosis of pro-thrombotic mediators including von Willebrand factor (vWF), angiopoietin-2 and P-selectin, all of which are elevated in the serum of severely affected patients and are significantly associated with in-hospital mortality [7-12]. Hospitalized patients also present with hyper-inflammation, having excessive levels of cytokines (cytokine storm) and other inflammatory markers that are characteristic of increased EC activity [13]. These data suggest that inappropriate degranulation of ECs could contribute to the pro-thrombotic and pro-inflammatory environment observed in severe COVID-19 cases. We therefore need to understand the molecular mechanisms underlying EC degranulation and determine how differences in this process confers susceptibility to the vascular abnormalities induced by COVID-19 and subsequent increased risk of death.

In ECs, pro-thrombotic and pro-inflammatory proteins are stored in specialized vesicles called Weibel-Palade bodies (WPBs) [14]. WPBs are endothelial cell-specific organelles that, upon release of contents, control important vascular events such as haemostasis, inflammation, angiogenesis and vascular tone. During vascular injury, WPBs undergo rapid degranulation and eject cargo into the vessel lumen including: vWF filaments to initiate platelet plug formation [15]; P-selectin to attract leukocytes [16] and angiopoietin-2 to induce EC migration [17]. WPBs also, in a positive feedback loop manner, store the chemokine CXCL8/ interleukin-8 [18]. Since the release of WPB cargo into the vessel lumen is highly regulated, there must be molecular changes that underlie the inappropriately raised plasma levels observed in COVID-19 patients. Excessive cargo release may be evoked by direct viral activation of ECs or an increase of cytokines acting on ECs, however the thrombo-inflammatory aberrations are not apparent in all infected patients (even those with high viral load). This indicates that there is either a pre-existing deterioration of the endothelium (co-morbidity) or patients may have an increased susceptibility to severe COVID-19 due to their genetic makeup, particularly in the genes that encode for proteins that play a role in the regulating the release of WPBs from ECs. Therefore, it is important to determine any links between variations in the genes that encode for proteins that regulate WPB cargo release with COVID-19 related mortality.

We previously reported a novel Rab GTPase (Rab46: CRACR2A-L; *Ensembl* CRACR2A-203; CRACR2A-a [19]) expressed in ECs that is necessary for anchoring a subpopulation of WPBs to the microtubule organising centre in response to histamine signalling [20]. This stimuli-coupled trafficking inhibits release of WPB cargo that are superfluous to histamine function and thus prevents the full thrombotic response evoked by vessel injury. Various mutations in this protein affect WPB cargo release in ECs, suggesting a regulatory role for Rab46 in WPB trafficking and cargo release. Rab46 (732 amino acids) is one of two functional isoforms from a possible six transcripts of the gene EFCAB4B and the only isoform expressed in ECs. CRACR2A (CRACR2A-S; *Ensembl* CRACR2a-201; CRACR2A-c), is a shorter (395 amino acids), non-Rab isoform of EFCAB4B expressed in T-cells and regulates store-operated calcium entry [21] and T-cell signalling [22] (Figure 1). Alignment of the amino acid sequences of Rab46 and CRACR2A variants reveals that the N-terminal components are identical, but Rab46 contains a distinct long C-terminus containing an extra Rab GTPase domain (Figure 1). Since an extreme cellular immune response is highlighted in patients with severe COVID-19, Rab46 and CRACR2A may also play an important role in the contribution of T-cell activity to disease severity. Indeed, we have reported that a patient with bi-allelic Rab46/CRACR2A mutations has a compromised immune system due to reduced T-cell signalling with subsequent defective cytokine production [23].

**Figure 1.**
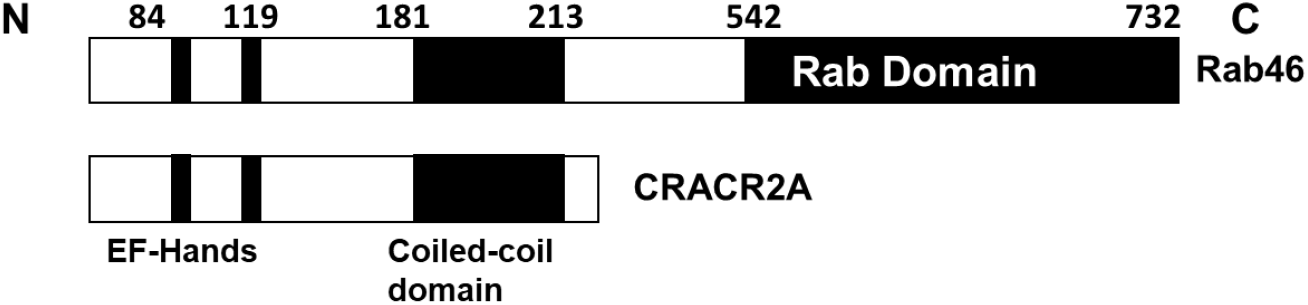
Schematics of the functional domains in the EFCAB4B isoforms Rab46 and CRACR2A

Recently, a longitudinal analysis of patients who recovered from mild COVID-19 reported decreased CRACR2A levels in the switch from an inflammatory to a wound healing response for the recovery phase of the infection [24]. Therefore, we hypothesise that genetic variations in EFCAB4B are associated with COVID-19 fatalities. Here, we used a candidate gene association study to determine the links between COVID-19 fatality and single nucleotide polymorphisms (SNPs) in the EFCAB4B gene using COVID-19 data recorded in the UK Biobank prior to the vaccination programme.

## Methods

### Demographic Data

The UK Biobank (application ID: 42651), from which we obtained data for use in this study, collected data on around 500,000 individuals in the UK between 2006-2010 [25]. These individuals consented to the UK Biobank obtaining their genomic data, health records and information regarding demographics and lifestyle. The UK Biobank released the data on 10,118 individuals prior to the vaccination programme, who self-identified as British and had been tested for COVID-19, with their associated outcomes also recorded. Co-morbidity data was also available for those individuals, including hypertension, diabetes, asthma, arthritis, stroke and myocardial infarction.

### Analysis of Rab46 SNPs associated with COVID-19 outcomes

Associations between Rab46 SNPs and COVID-19 fatality were determined by logistic regression analysis, using an additive method. The genotype data were mainly handled with the PLINK 2.0 analytical framework [26]. The initial screening process generated the samples from British ethnic background (coding: 1001) that have COVID-19 test data and the variants with imputation info score > 0.4. Further filtering was carried out using --geno 0.1, --maf 0.01, and --hwe 0.000001 for variants and using --mind 0.1 for samples. In addition, only the variants residing on EFCAB4B gene have been retained according to start coordinate (3714799) and end coordinate (3874985) on human chromosome 12 based on the genome assembly version GRCh37. The three cohorts were defined as follows. Fatal group comprises individuals who tested positive for COVID-19 and who are also labelled with COVID-19 deaths (coding: U071). Non-fatal group consists of individuals who tested positive for COVID-19 with no records in the cause of death data. Negative group is made up of those who tested negative for COVID-19 that are not identified in the cause of death data. To test association of genotypes with phenotypes, the logistic regression was performed for case/control phenotypes accounting for covariates including sex, age and the first ten principal components. This enables identification of whether the presence of SNP is a risk factor for COVID-19 fatality by comparing the three cohorts (fatal vs negative, fatal vs non-fatal and non-fatal vs negative). For the adjustment of illnesses, additional covariate has been added respectively for the six comorbidities such as hypertension, myocardial infarction, stroke, diabetes mellitus, arthritis and asthma obtained from non-cancer illness code. Ensembl Variant Effect Predictor (VEP) was used to predict and annotate the effect and impact of the variant on the gene functions from the human genome (GRCh37). The SNPs identified in Table 2 were those that caused missense mutations and were statistically significant (P < 0.05).

### Conservation of Rab46 protein sequence across different species

Rab46 sequence conservation diagram was generated using *Basic Local Alignment Search Tool* (BLAST; https://blast.ncbi.nlm.nih.gov/Blast.cgi) and WebLogo 3 (http://weblogo.threeplusone.com). Amino acid sequence of Rab46 (NP_001138430.1) was searched against protein sequence database in BLAST (BLASTP) and aligned with the identified 100 similar protein sequences (percent sequence identity > 80 %) from 66 different species using COBALT multiple alignment tool. The generated sequence alignment enabled calculation of residue conservation scores. WebLogo 3 [27, 28] was used to visualise the level of amino acid sequence conservation around residues for which missense mutations were detected.

### Rab46 structural analysis

The predicted AlphaFold [29, 30] structure of full-length human Rab46 (https://alphafold.ebi.ac.uk/entry/Q9BSW2) and the crystal structure of EF-hand domain (PDB: 6PSD) were pre-processed and energy-minimised using the Protein Preparation Wizard (Maestro software, Release 2020-1, Glide, Schrödinger, LLC, New York, NY, 2020).

The effect of the A98T mutation was modelled using the available crystal structure of EF-hand domain of Rab46. Alanine residue at position 98 was mutated to threonine using ‘Mutate Residue’ tool in the Maestro graphical user interface (Maestro software, Release 2020-1, Glide, Schrödinger, LLC, New York, NY, 2020). Side chains of amino acids within 5 Å from the introduced mutation were minimised using the OPLS3 forcefield [31]. Alignment of the wild type and mutant EF-hand domain structures was visualised using PyMOL (The PyMOL Molecular Graphics System, Version 2.0 Schrödinger, LLC.) and changes to the structure visualised using Pymol and the Maestro graphical user interface.

## Results

### Demographics

Here we analyzed genomic data (exonic and intronic) held in the UK Biobank from 10,118 subjects, who had self-identified as British. Of these 51% were female and 49% males, with a mean age of 69 ± 8.3 years (means ± SD). In total, 1265 patients were diagnosed COVID-19 positive (Table 1). The positive cases were further categorised into fatal and non-fatal, with fatal indicating the individuals who died due to COVID-19 based on data obtained from the National Death Registries Database. Among those who tested positive, 194 (15.3%) were reported to have died due to COVID-19 (fatal), while 1071 (84.7%) were non-fatal. In addition, because any determined associations between EFCAB4B SNPs and COVID19 fatality could be due to the SNP predisposing an individual to a COVID-19 susceptible health condition, we evaluated the co-morbidities of these individuals. Of the 1265 COVID-19-positive individuals, 426 (33%) self-reported hypertension, 54 (4.27%) myocardial infarction, 35 (2.77%) stroke, 104 (8.22%) diabetes mellitus, 152 (12%) arthritis and 158 (12.5 %) asthma.

**Table 1.** Demographics of UK Biobank patients used in this study. N = the number of individuals tested for COVID-19 in the UK Biobank

| Characteristic | N | Covid-19 negative | Covid-19 positive | Fatal | Non-Fatal |
| --- | --- | --- | --- | --- | --- |
| *Number of individuals* | 10118 | 8853 | 1265 | 194 | 1071 |
| *Age* | 69 ± 8.3 | 70.1 ± 8.2 | 70.4 ± 7.5 | 75.1 ± 5.7 | 67.8 ± 9.2 |
| **Gender** |  |  |  |  |  |
| *Male* | 4972 | 4309 | 663 | 129 | 534 |
| *Female* | 5146 | 4544 | 602 | 65 | 537 |
| **Co-Morbidity** |  |  |  |  |  |
| *Myocardial Infarction* | 429 | 375 | 54 | 13 | 41 |
| *Stroke* | 256 | 221 | 35 | 6 | 29 |
| *Arthritis* | 1477 | 1325 | 152 | 37 | 115 |
| *Diabetes Mellitus* | 731 | 627 | 104 | 27 | 77 |
| *Hypertension* | 3436 | 3010 | 426 | 86 | 340 |
| *Asthma* | 1380 | 1222 | 158 | 17 | 141 |

Using logistic regression analysis, a candidate gene association study was performed to explore potential links between COVID-19 fatality and SNPs found in the EFCAB4B gene. Association between known SNPs in EFCAB4B gene and COVID-19 outcomes were analysed by calculating Odds Ratios (ORs) from pairwise analysis of the populations described in Table 1. We created three comparative cohorts:-1) Non-fatal vs negative: Patients who tested positive for COVID-19 and survived (case) as compared to those who had not been diagnosed with COVID-19 (control): 2) Fatal vs negative: Patients who tested positive for COVID-19 and died (case) as compared to those who had not been diagnosed with COVID-19 (control): 3) Fatal vs non-fatal: Patients who had tested positive for COVID-19 and died (case) as compared to those who survived (control). Figure 2 displays the distribution of all the variants (exonic and intronic) significantly (p = < 0.05) associated with the test cases in each comparative cohort. A total of 127 variants are associated with COVID-19 infection (both fatal and non-fatal) compared with non-infected population, 104 variants were associated with COVID-19 fatality as compared to non-infected populations and from these, 53 variants are significantly associated with fatality compared to non-fatal cases. This suggests that overrepresentation of these 53 variants is not due to disease susceptibility. Herein, we specifically explored SNPs that affect translation of the EFCAB4B gene into its relevant proteins by inducing missense mutations. Three SNPs with a minor allele frequency (MAF) > 0.05 were identified as causing missense mutations in both CRACR2A and Rab46 (Table 2). We determined these SNPs as being significantly associated with COVID-19 fatality in the fatal vs negative cohort as compared to the non-fatal group (Table 3). The results showed that: 1) the C allele of rs9788233 is significantly associated with mortality with an OR of 1.51 (p = 0.004): 2) the T allele of rs17836273 has an OR of 1.43 (p = 0.012): 3) the T allele of rs36030417 has an OR of 1.39 (p = 0.013) (Table 2). Since COVID-19 morbidity may not be directly associated with the effects of EFCAB4B variant but by covariant effects, we thus explored whether these missense variants reflect true associations between Rab46/CRACR2A and COVID-19 fatality and adjusted the data to account for six underlying disorders: hypertension, myocardial infarction, stroke, diabetes mellitus, arthritis and asthma (Table 4). Consistently across six models of logistic regression, the results supported relevance of the same three missense variants to COVID-19 fatality.

**Figure 2.**
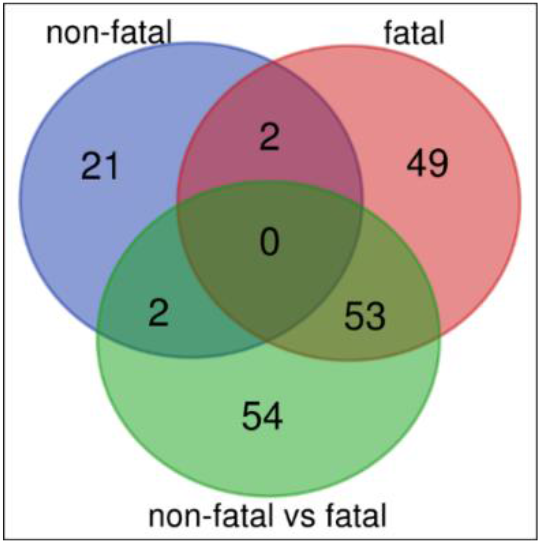
Venn diagram displaying the distribution of exonic and intronic SNPs associated with the test cases in each of the three cohorts: non-fatal (non-fatal vs negative), fatal (fatal vs negative) and non-fatal vs fatal.

**Table 2.** Characterisation of three missense Rab46/CRACR2A SNPS. MAF = mean allele frequency (ALFA project [2]). REF = reference allele. ALT = alternative allele

| SNIP ID | MAF (alfa) | REF | ALT | TYPE | Amino acid change |
| --- | --- | --- | --- | --- | --- |
| rs9788233 | 0.118 | T | C | substitution | R7G |
| rs17836273 | 0.123 | C | T | substitution | A98T |
| rs36030417 | 0.1453 | A | T | substitution | H212Q |

**Table 3.** Association analysis of three missense Rab46/ CRACR2A SNPS with COVID-19 outcomes. OR = Odds Ratio. Non-fatal = Non-fatal vs negative cohort. Fatal = Fatal vs negative cohort. Fatal (vs non) = Fatal vs non-fatal cohort. – indicates no significant variants found.

| SNP ID | P Value/ OR | P Value/ OR | P Value/ OR |
| --- | --- | --- | --- |
|  | Non-fatal | Fatal | Fatal (vs non) |
| rs9788233 | - | 0.004 / 1.511 | 0.021 / 1.486 |
| rs17836273 | - | 0.012 / 1.433 | 0.099 / 1.340 |
| rs36030417 | - | 0.013 / 1.393 | 0.055 / 1.350 |

**Table 4.** Covid-19 outcome and Rab46 missense SNPs association analysis after adjustment for co-morbidities. OR = odds ratio

| Patient Cohort comparison | SNP ID | Hypertension<br>P / OR | Diabetes<br>P / OR | Asthma<br>P / OR | Arthritis<br>P / OR | Stroke<br>P / OR | Myocardial infarction<br>P / OR |
| --- | --- | --- | --- | --- | --- | --- | --- |
| Fatal Vs Negative | rs9788233 | 0.004 / 1.51 | 0.004 / 1.51 | 0.004 / 1.51 | 0.004 / 1.51 | 0.004 / 1.51 | 0.004 / 1.51 |
|  | rs17836273 | 0.012 / 1.44 | 0.012 / 1.44 | 0.014 / 1.43 | 0.012 / 1.43 | 0.012 / 1.43 | 0.012 / 1.43 |
|  | rs36030417 | 0.013 / 1.39 | 0.013 / 1.4 | 0.014 / 1.39 | 0.013 / 1.39 | 0.013 / 1.39 | 0.013 / 1.39 |
| Fatal Vs Non-fatal | rs9788233 | 0.022 / 1.48 | 0.018 / 1.5 | 0.024 / 1.48 | 0.024 / 1.47 | 0.022 / 1.48 | 0.021 / 1.49 |
|  | rs17836273 | 0.105 / 1.31 | 0.089 / 1.33 | 0.102 / 1.31 | 0.12 / 1.30 | 0.104 / 1.31 | 0.099 / 1.31 |
|  | rs36030417 | 0.013 / 1.39 | 0.050 / 1.36 | 0.056 / 1.35 | 0.062 / 1.35 | 0.053 / 1.35 | 0.055 / 1.35 |

These three missense SNPs induce amino acid substitutions in both CRACR2A and Rab46 protein when translated from EFCAB4B (Table 2 and Figure 3). The arginine to glycine (R7G) change induced by rs9788233 occurs at the N-terminus in a structurally undefined region. The substitutions incurred by both rs17836273 and rs36030417 (alanine to threonine A98T and histidine to glutamine H212Q respectively) occur in functionally important domains found in both CRACR2A and Rab46 (the canonical motif of the EF-hand domain and the coiled-coil domain). In order to determine the importance of these amino acid residues we explored their evolutionary conservation (Figure 4). The conservation of Rab46 across 100 different sequences and 66 species was obtained through conducting BLAST sequence alignments of the Rab46 protein sequence. From the multiple alignments, it was found that arginine (rs9788233) in a domain of unknown function was conserved across 80% of species. Other common amino acids found in the 20% of non-conserved species were lysine and asparagine. Alanine (rs17836273) at position 98 in the second EF-hand motif was highly conserved (98%) and histidine (rs36030417) located in the coiled-coil domain was the most conserved (99%). The high level of conservation of these residues suggest that these amino acids are critical to the function of Rab46 and CRACR2A.

**Figure 3.**
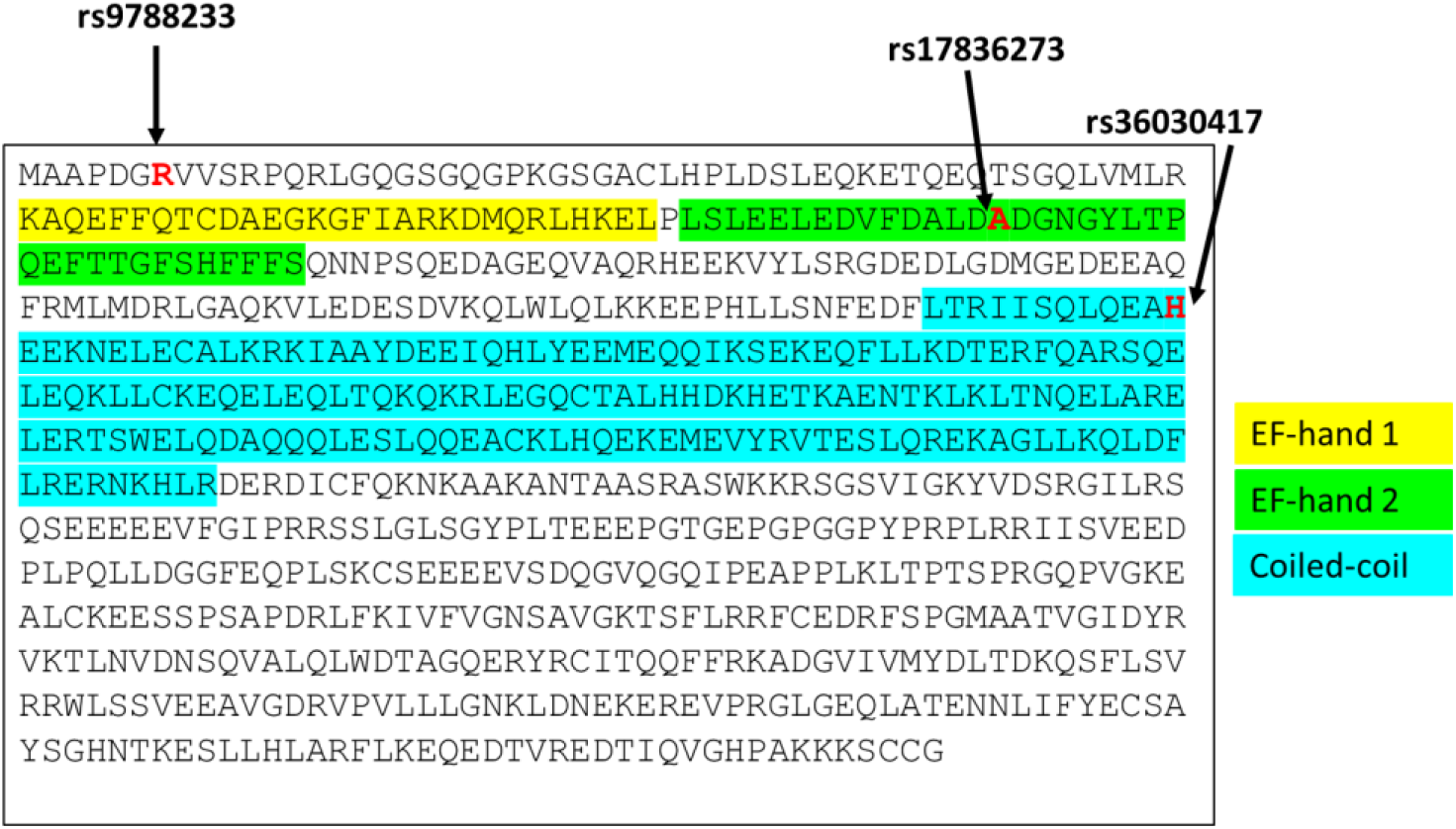
Human amino acid sequence of Rab46 indicating the position of the SNPs (highlighted in red) significantly associated with COVID-19 fatality within the highlighted functional domains.

**Figure 4.**
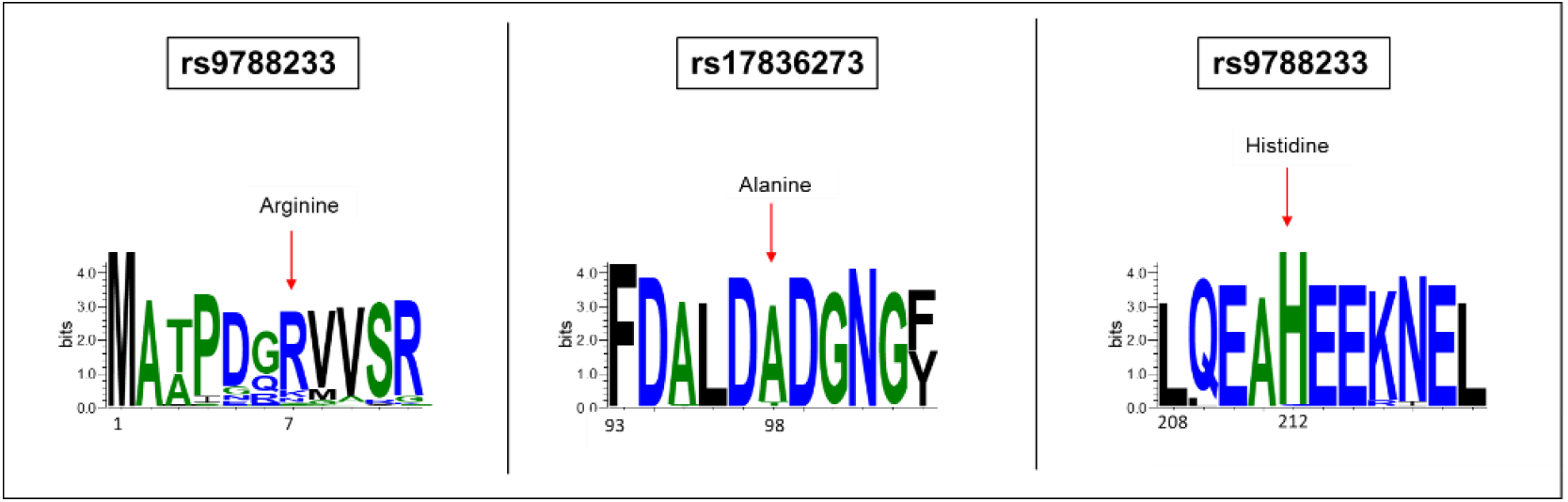
Sequence logo plots depicting the conservation of the specified amino acid residues compared across 100 sequences and 66 species demonstrates the high conservation of the three SNPs

The structure of Rab46 is unknown but we sought further insight into the role of the SNPs in protein function by analysis of an AlphaFold prediction of the structure of full-length Rab46 and the available crystal structure of the EF-hand domain (Figure 5). The AlphaFold prediction of the EF-hand and the coiled-coil domains are of high confidence, and here we suggest that these residues have outward facing side chains (highlighted in yellow).

**Figure 5.**
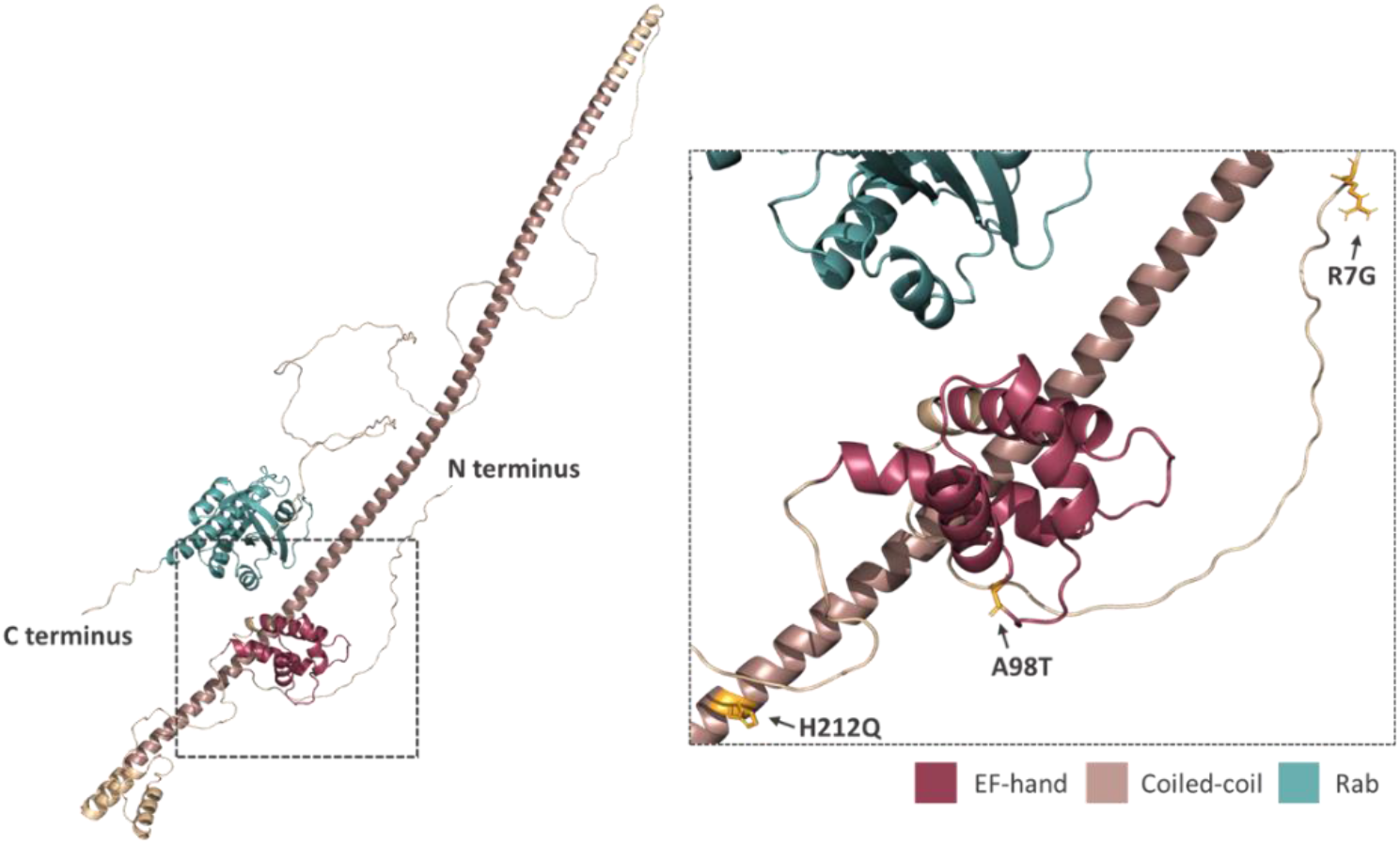
Prediction of the position and orientation of the three missense SNPs in an AlphaFold model of full-length Rab46 structure. The box indicated by the dotted line in the full-length structure is enlarged in order to demonstrate the position and orientation of the side chains of the three missense SNPs: R7G; A98T; H212Q.

We further explored the A98T mutation through the availability of the crystal structure for the EF-hand domain (PDB: 6PSD). To understand the possible impact of the threonine residue that is substituted for alanine (A98T) in rs17836273 we used molecular modelling to overlay the crystal structure of the wild-type (WT) alanine-containing variant (blue) with the threonine substituted variant (cream: Figure 6). Rab46 contains two EF-hand motifs, however the crystal structure and Ca^2+^ binding data predict that only the second EF-hand coordinates Ca^2+^ (Figure 6). The aspartate/alanine/aspartate (DAD: the WT structure) motif in this second EF-hand is necessary for coordinating calcium [32]. Overlaying the WT crystal structure with the predicted A98T variant (shown in cream) reveals small structural changes to the binding domain where the coordinating aspartic acid residues become juxtaposed when overlaid, suggesting this variant could cause deficiencies in Ca^2+^ coordination and protein conformation.

**Figure 6.**
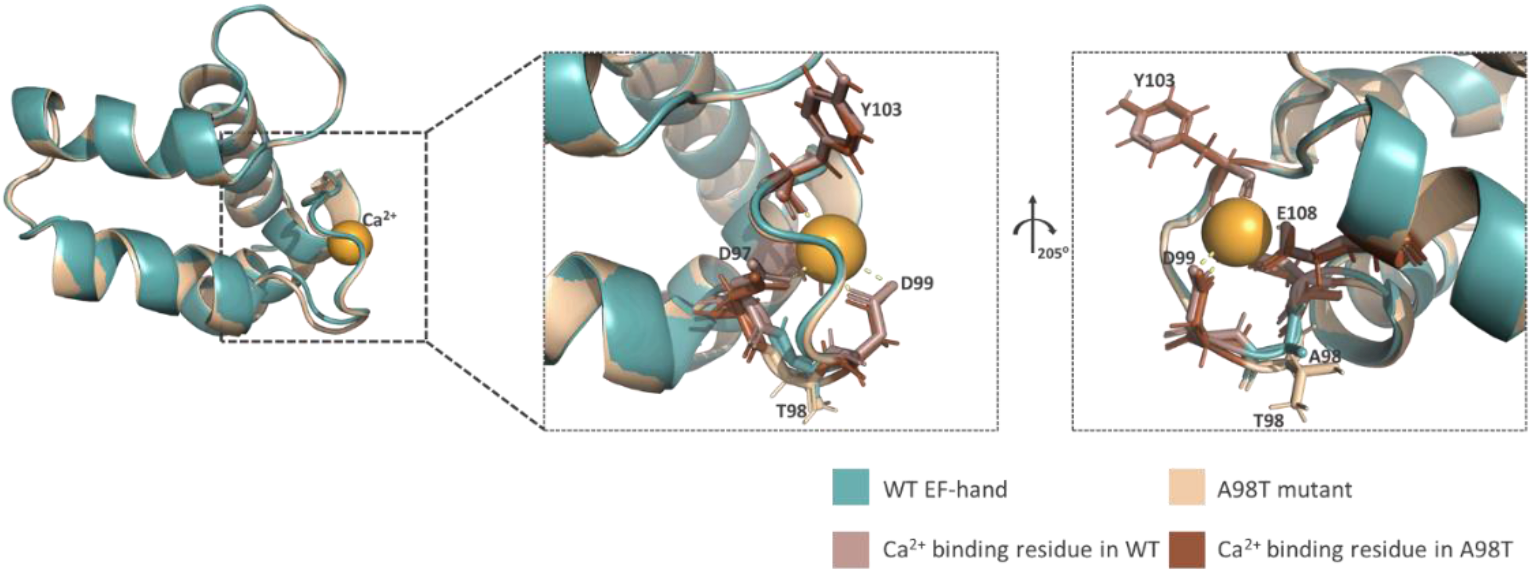
Modelling of the effect of A98T substitution on the EF-hand domain of Rab46. Right: crystal structure of the wild type (WT) and a modelled structure of the mutant (A98T) EF-hand domain aligned using PyMOL. Left: expanded boxes demonstrating subtle changes within the calcium binding loop.

In summary, we have described three missense mutations in EFCAB4B gene that are associated with fatality in patients with COVID-19, of which two could cause changes to protein structure and function.

## Discussion

Here we have performed logistic regression analysis using recorded COVID-19 outcomes from patient data deposited in the UK Biobank, prior to the vaccination programme, and links between known EFCAB4B SNPs. No missense SNPs were associated with non-fatal COVID-19 but three missense mutations that caused substitutions in the translated protein were significantly associated with fatality. The data suggests that these SNPs are not linked to susceptibility of infection with COVID-19 but that genetic variation in EFCAB4B gene is a factor in COVID-19 fatality.

Two missense mutations, rs17836273 and rs36030417, are highly conserved (98% and 99% respectively) and are located in the N-terminus of both CRACR2A and Rab46 protein in domains that have roles in protein function. Mutation rs36030417 causes a histidine to glutamine switch in a region that is predicted to be a coiled-coil domain. Coiled-coils have several important roles in proteins but they are mostly important for facilitating protein-protein interactions. Histidine and glutamine are fairly conserved in size, which is important for residues that have to be packed inside an alpha helix, however the positively charged imidazole ring on histidine could be important for metal coordination which is often used in coiled-coils for reinforcement of an oligomerized protein complex [33]. Particularly here, it is suggested by the predicted AlphaFold structure that the amino acid at position 212 orientates outwardly and therefore would be more important in molecular interactions than helix stability. Both Rab46 and CRACR2A are suggested to act as multimers and therefore mutation and biochemical analysis of this variant is necessary to determine the impact it has on this function.

Mutation rs17836273 is located in the EF-hand domain of both Rab46 and CRACR2A. EF-hands consist of a helix-loop-helix motif and binding of calcium to the loop region causes a change in the EF-hand conformation, that is propagated through a protein to enable function. The second EF-hand of Rab46 consists of a canonical calcium binding motif of 12 amino acids (DADGNGYLTPQE). Amino acids in position 1, 3, 5, 7, 9 and 12 in this motif are important in the coordination of the calcium ion [34]. The A98T substitution conferred by this SNP is a fairly conserved alanine to threonine mutation and it is located in position 2 of the motif, which is thought to be a changeable site. However, using the crystal structure of the EF-hand domain and molecular modelling to overlay the two variants we suggest that the shape of the calcium binding loop is slightly altered by the threonine substitution, where the negatively charged aspartic acid residues that coordinate the positively charged calcium ion are shifted so that they look juxtaposed when overlaid. Although the model suggests that calcium is able to bind to the loop, we predict that these conformational differences evoke changes in protein function that, although tolerated within a population (the SNP occurs in 12.3% of the overall population), will overtime or under highly stressful conditions (e.g. COVID-19), have a detrimental effect. We do know that alanine to threonine substitutions in disease causing proteins such as amyloid display antagonistic pleiotrophism [35], which are mutations that whilst advantageous when young exert problems in the aging population, which explains why these polymorphisms are preserved within a population. The effect of A98T will need further biochemical analysis to determine the impact on both calcium binding and cell physiology.

Similar coiled-coil and EF-hand-containing proteins (e.g. STIM1) have mutations in these domains that cause disease, for example an activating mutation in STIM1 coiled-coil region is associated with Stormoken syndrome [36] whilst an EF-hand mutation causes tubular aggregate myopathy [37]. We do not know if these variants in CRACR2A/Rab46 evoke activation or inactivation. We know that in endothelial cells, Rab46 acts as a brake in response to certain stimuli to prevent total release of WPB contents [20]. Therefore, we could predict that inhibition of this function would lead to an increase in the release of pro-thrombotic and pro-inflammatory cargo thus leading to some of the clinical features observed in severe cases of COVID-19. In T-cells Rab46 is important for T-cell signalling [22], moreover we have shown that a patient who has biallelic mutations in the gene EFCAB4B (double mutation in allele1: R144G and E300*; allele 2 E278D), so that they have no longer express full length Rab46 or CRACR2A, exhibits reduced cytokine expression due to decreased calcium influx and JNK signalling [23]. This suggests that overexpression or over-activation of Rab46 in T-cells could lead to the increase in cytokine release as observed in patients with cytokine storm reactions to COVID-19.

A role for Rab46/CRACR2A in the immune response to COVID-19 infection is indicated in a study by Furuyama et al. [38]. Intramuscular injection of a SARS CoV-2 spike-containing vaccine in Rhesus macaques resulted in a cellular and humoral immune response and provided protection from COVID-19 driven pneumonia compared to intranasal administration. Transcriptional analysis of differentially expressed genes from lung tissues of infected animals demonstrated upregulation of Rab46/CRACR2A in the protected monkeys. Moreover, in a study by Talla et al, characterisation of signals associated with recovery in individuals with mild COVID-19 demonstrated a reduced expression of inflammatory proteins, including Rab46/CRACR2A, after the early immune response [24]. These studies indicate that regulating the expression of Rab46 and/or CRACR2A is important for both the initial response and recovery phases of COVID-19 infection.

To understand the importance of the EFCAB4B missense variations identified as being associated with COVID-19 fatality requires further structural analysis and longer term studies using appropriate animal models. However, we also need to be aware of the limitations of the study because the reported data are from a relatively small sample size in an aged population prior to the rollout of a vaccination programme in the UK. In addition, we do not have a defined list of all the comorbidities that are associated with COVID-19 susceptibility. Here, we have also focussed on the variations that culminate in missense mutations, however, many of the intronic variations identified as being significantly associated with COVID-19 fatality could be of interest especially if they cause changes in expression levels or isoform splicing.

## Data Availability

All data produced in this present study are available upon reasonable request to the authors

## Acknowledgements

This research has been conducted using the UK Biobank Resource (Project ID: 42651). The work was supported by a Medical Research Council grant to LM (<u>MR/T004134/1</u>). The study involved use of ARC4, which is part of the High Performance Computing Facility at the University of Leeds. We’d like to thank LeedsOmics for all their input (https://omics.leeds.ac.uk/).

## Author Contributions

DW and CWC performed genetic analysis. SDW and AC performed the conservation analysis. SDW and KJS performed molecular modelling and ALB advised on interpretation of the AlphaFold model and crystal structures. LP, AC and AM analysed the data and created the tables. LM conceived the study and co-wrote the article with input from all authors. LM generated research funds and coordinated the project.

## References

1. Atzrodt, C.L., et al., A Guide to COVID-19: a global pandemic caused by the novel coronavirus SARS-CoV-2. Febs j, 2020. 287(17): p. 3633–3650.

2. L. Phan, Y.J., H. Zhang, W. Qiang, E. Shekhtman, D. Shao, D. Revoe, R. Villamarin, E. Ivanchenko, M. Kimura, Z. Y. Wang, L. Hao, N. Sharopova, M. Bihan, A. Sturcke, M. Lee, N. Popova, W. Wu, C. Bastiani, M. Ward, J. B. Holmes, V. Lyoshin, K. Kaur, E. Moyer, M. Feolo, and B. L. Kattman. ALFA: Allele Frequency Aggregator. www.ncbi.nlm.nih.gov/snp/docs/gsr/alfa/ 2020.

3. McFadyen, J.D., H. Stevens, and K. Peter, The Emerging Threat of (Micro)Thrombosis in COVID-19 and Its Therapeutic Implications. Circ Res, 2020. 127(4): p. 571–587.

4. Wazny, V., et al., Vascular underpinning of COVID-19. Open Biol, 2020. 10(8): p. 200208.

5. Nägele, M.P., et al., Endothelial dysfunction in COVID-19: Current findings and therapeutic implications. Atherosclerosis, 2020. 314: p. 58–62.

6. Zhang, J., K.M. Tecson, and P.A. McCullough, Endothelial dysfunction contributes to COVID-19-associated vascular inflammation and coagulopathy. Rev Cardiovasc Med, 2020. 21(3): p. 315–319.

7. Goshua, G., et al., Endotheliopathy in COVID-19-associated coagulopathy: evidence from a single-centre, cross-sectional study. Lancet Haematol, 2020. 7(8): p. e575–e582.

8. Villa, E., et al., Dynamic angiopoietin-2 assessment predicts survival and chronic course in hospitalized patients with COVID-19. Blood Adv, 2021. 5(3): p. 662–673.

9. Fenyves, B.G., et al., Plasma P-selectin is an early marker of thromboembolism in COVID-19. Am J Hematol, 2021. 96(12): p. E468–e471.

10. Yatim, N., et al., Platelet activation in critically ill COVID-19 patients. Ann Intensive Care, 2021. 11(1): p. 113.

11. Cotter, A.H., et al., Elevated von Willebrand Factor Antigen Is an Early Predictor of Mortality and Prolonged Length of Stay for Coronavirus Disease 2019 (COVID-19) Inpatients. Arch Pathol Lab Med, 2022. 146(1): p. 34–37.

12. Philippe, A., et al., Circulating Von Willebrand factor and high molecular weight multimers as markers of endothelial injury predict COVID-19 in-hospital mortality. Angiogenesis, 2021. 24(3): p. 505–517.

13. Hu, B., S. Huang, and L. Yin, The cytokine storm and COVID-19. J Med Virol, 2021. 93(1): p. 250–256.

14. Naß, J., J. Terglane, and V. Gerke, Weibel Palade Bodies: Unique Secretory Organelles of Endothelial Cells that Control Blood Vessel Homeostasis. Front Cell Dev Biol, 2021. 9: p. 813995.

15. Karampini, E., R. Bierings, and J. Voorberg, Orchestration of Primary Hemostasis by Platelet and Endothelial Lysosome-Related Organelles. Arterioscler Thromb Vasc Biol, 2020. 40(6): p. 1441–1453.

16. Kawecki, C., P.J. Lenting, and C.V. Denis, von Willebrand factor and inflammation. J Thromb Haemost, 2017. 15(7): p. 1285–1294.

17. Cossutta, M., et al., Weibel-Palade Bodies Orchestrate Pericytes During Angiogenesis. Arterioscler Thromb Vasc Biol, 2019. 39(9): p. 1843–1858.

18. Wolff, B., et al., Endothelial cell “memory” of inflammatory stimulation: human venular endothelial cells store interleukin 8 in Weibel-Palade bodies. J Exp Med, 1998. 188(9): p. 1757–62.

19. Wilson, L.A., et al., Expression of a long variant of CRACR2A that belongs to the Rab GTPase protein family in endothelial cells. Biochem Biophys Res Commun, 2015. 456(1): p. 398–402.

20. Miteva, K.T., et al., Rab46 integrates Ca(2+) and histamine signaling to regulate selective cargo release from Weibel-Palade bodies. J Cell Biol, 2019. 218(7): p. 2232–2246.

21. Srikanth, S., et al., A novel EF-hand protein, CRACR2A, is a cytosolic Ca2+ sensor that stabilizes CRAC channels in T cells. Nat Cell Biol, 2010. 12(5): p. 436–46.

22. Woo, J.S., et al., CRACR2A-Mediated TCR Signaling Promotes Local Effector Th1 and Th17 Responses. J Immunol, 2018. 201(4): p. 1174–1185.

23. Wu, B., et al., Biallelic mutations in calcium release activated channel regulator 2A (CRACR2A) cause a primary immunodeficiency disorder. Elife, 2021. 10.

24. Talla, A., et al., Longitudinal immune dynamics of mild COVID-19 define signatures of recovery and persistence. bioRxiv, 2021: p. 2021.05.26.442666.

25. Bycroft, C., et al., The UK Biobank resource with deep phenotyping and genomic data. Nature, 2018. 562(7726): p. 203–209.

26. Chang, C.C., et al., Second-generation PLINK: rising to the challenge of larger and richer datasets. Gigascience, 2015. 4: p. 7.

27. Crooks, G.E., et al., WebLogo: a sequence logo generator. Genome Res, 2004. 14(6): p. 1188–90.

28. Schneider, T.D. and R.M. Stephens, Sequence logos: a new way to display consensus sequences. Nucleic Acids Res, 1990. 18(20): p. 6097–100.

29. Jumper, J., et al., Highly accurate protein structure prediction with AlphaFold. Nature, 2021. 596(7873): p. 583–589.

30. Varadi, M., et al., AlphaFold Protein Structure Database: massively expanding the structural coverage of protein-sequence space with high-accuracy models. Nucleic Acids Res, 2022. 50(D1): p. D439–d444.

31. Harder, E., et al., OPLS3: A Force Field Providing Broad Coverage of Drug-like Small Molecules and Proteins. J Chem Theory Comput, 2016. 12(1): p. 281–96.

32. Lee, I.G., et al., A tunable LIC1-adaptor interaction modulates dynein activity in a cargo-specific manner. Nat Commun, 2020. 11(1): p. 5695.

33. Boyle, A.L., et al., Selective coordination of three transition metal ions within a coiled-coil peptide scaffold. Chemical Science, 2019. 10(31): p. 7456–7465.

34. Gifford, J.L., M.P. Walsh, and H.J. Vogel, Structures and metal-ion-binding properties of the Ca2+-binding helix-loop-helix EF-hand motifs. Biochem J, 2007. 405(2): p. 199–221.

35. Podoly, E., G. Hanin, and H. Soreq, Alanine-to-threonine substitutions and amyloid diseases: butyrylcholinesterase as a case study. Chem Biol Interact, 2010. 187(1-3): p. 64–71.

36. Misceo, D., et al., A dominant STIM1 mutation causes Stormorken syndrome. Hum Mutat, 2014. 35(5): p. 556–64.

37. Noury, J.B., et al., Tubular aggregate myopathy with features of Stormorken disease due to a new STIM1 mutation. Neuromuscul Disord, 2017. 27(1): p. 78–82.

38. Furuyama, W., et al., Rapid protection from COVID-19 in nonhuman primates vaccinated intramuscularly but not intranasally with a single dose of a recombinant vaccine. bioRxiv, 2021: p. 2021.01.19.426885.

